# CNA Explorer and anaLyzer (CNAEL): an interactive web application and standard operating procedure enabling efficient clinical review and reporting of complex NGS-derived tumor copy number profiles

**DOI:** 10.1101/2022.10.24.22281236

**Authors:** Ellen Chen, Jinlian Wang, Robert Kueffner, Hussam Al-Kateb, Antonina Silkov, Andrew Uzilov, Lucas Lochovsky, Hui Li, Scott Newman

## Abstract

**Purpose:** Clinical analysis and reporting of somatically acquired copy number abnormalities (CNAs) detected through next-generation sequencing (NGS) is time consuming and requires significant expertise. Interpretation is complicated by other classes of variants such as coding mutations and gene fusions. Recent guidelines for the clinical assessment of tumor CNAs harmonize and simplify the reporting criteria but did not directly address NGS-specific concerns or the need for a standardized and scalable protocol for CNA analysis.

**Methods:** We developed a scalable NGS-derived CNA analysis protocol paired with a novel interactive web application, CNA Explorer and anaLyzer (CNAEL), to facilitate the rapid, scalable, and reproducible analysis and reporting of complex tumor-derived CNA profiles https://CNAEL.sema4.com.

**Results:** Novel features of CNAEL include on-the-fly data rescaling to account for tumor ploidy, purity, and modal chromosomal copy number; integration of gene expression, coding, and fusion variants into review and automated genome-wide summarization to enable rapid reporting. We found that case curation times were significantly reduced when using CNAEL [median:7 mins, IQR = 4, 10.25] compared with our previous laboratory standard operating procedure [median: 61 mins, IQR = 23.75, 176,25] with p=4.631e-05.

**Conclusion:** CNAEL enables efficient and accurate clinical review and reporting of complex NGS-derived tumor copy number profiles.

## Introduction

Copy number abnormalities and loss of heterozygosity (CNAs; LOH) acquired by tumors are important diagnostic, prognostic, and therapeutically relevant features of tumor genomes. For example, 1p/19q codeletion is pathognomonic for oligodendroglioma ^1^, hyperdiploidy (gain of multiple chromosomes) in acute lymphoblastic leukemia, which defines a distinct prognostic group ^2^ and amplified *ERBB2* in breast cancer that can be targeted with specific inhibitors ^3^. As such, copy number analysis forms an integral part of clinical tumor profiling: either through karyotyping, fluorescence *in situ* hybridization (FISH), copy number microarray or more recently, next generation sequencing (NGS) approaches, including whole genome and whole exome ^4,5^.

Interpretation of genome-wide copy number segmentation profiles is challenging. Cancer genomes can be highly unstable and vary widely in the number of bioinformatically called genomic segments, copy number states, segment sizes, and gene content. Further complications include whole genome duplication and polyploidization, intratumor heterogeneity, the presence of non-tumor tissue, and false positive and negative CNA calls due to bioinformatic over/under-segmentation ^6,7^. Therefore, clinical review of copy number profiles is complex and highly nuanced, requiring a high level of subject matter expertise. Review is also labor intensive because historically, profiles have been analyzed and reported segment by segment with each being assessed for cancer relevant gene content. Thus, clinical copy number reports are laborious to produce; they can contain hundreds of copy number segments and are often challenging for oncologists to interpret ^8^.

In recognition of the clinical value of CNAs and the complexity of the analysis process, a joint Association for American College of Medical Genetics and Genomics (ACMG) and the Cancer Genomics Consortium (CGC) developed consensus guidelines for the clinical reporting of CNAs ^9^. Important innovations include a well-defined tier system to guide reporting and to prioritize clinically significant variants on reports, the ability to summarize complex and chromothripsis regions in a single call, and the recommendation to report significant patterns of abnormalities as a single entity. While these guidelines represent a huge step forward, they do not directly address many of the complications listed above, nor do they focus on the operational challenges or opportunities presented by newer WES/WGS and multiplatform NGS approaches. Specific challenges are: 1). how to efficiently operationalize the copy number analysis workflow; 2). how to leverage NGS data to produce meaningful numerical copy number estimates rather than a less interpretable copy number ratio; and 3). how to integrate CNA with sequence-level and structural mutations into a holistic genome-wide analysis.

To address these challenges and opportunities, we developed an efficient five-step CNA analysis Standard Operating Procedure (SOP) that encompasses key recommendations of the ACMG/CGC guidelines accompanied by an interactive web application, CNA Explorer and anaLyzer (CNAEL), which facilitates the SOP. Key features of CNAEL are: i) an ability to perform on-the-fly rescaling of copy number probes and segments to account for tumor purity and ploidy; ii) intrinsic support for multi-modal analysis by superimposing sequence level SNV/INDEL mutations, gene fusions, and gene expression outliers on the CNV profile; iii) summarization of complex and chromothripsis regions; iv) auto-summarization of complex copy number profiles to enable rapid reporting; and iv) knowledgebase integration to assist in efficient review. Lastly, we assessed the efficiency and accuracy of the SOP and web appplication by reviewing a series of challenging cases from our clinical service and show that we significantly shorten the analysis time without sacrificing accuracy.

## Materials and Methods

### Example case selection

We randomly selected twenty cases that represent 17 distinct OncoTree codes from our clinical NGS service. We noted that our service focuses on formalin-fixed paraffin-embedded solid tumors and all cases were de-identified prior to analysis. The study did not directly involve human subjects, and the software is designed for laboratory quality assurance and process optimization; therefore, IRB was not required.

### Exome data and bioinformatics processing

Illumina NovaSeq 100 basepair paired-end sequencing was performed in our CLIA lab using Twist Core Exome capture to a target coverage of 250X for tumor and 100X for normal samples. We aligned FASTQ pairs to the human_g1k_v37 reference genome using the Broad Institute’s GATK4.0 best practices. The alignment was performed with BWA-MEM, duplicate marking by Picard, and base quality score recalibration with GATK4 version 4.2.0.0. We used tumor and normal BAM files to generate copy number segmentation .seg files, which followed the Broad institute’s GATK4.0 best practices pipeline^10^.

### Description of the web application

CNAEL is a web-based tool that supports somatic CNA visualization, rescaling, correction, editing, and profile rescaling. The tool accepts user-supplied file inputs which includes: the genomic coordinates and copy number calls, copy number probes, somatic variants, germline variants, fusions, and RNA expression data. The public version supports user-supplied data files which can be uploaded via web interface.

### User-generated input data

1. ”ProbeData.seg” file containing probe-level data. In the case of exome sequencing, each probe coincided with a captured exon. For WGS, genome regions are typically binned into regions of ∼100 basepairs. Probe-level data is in Integrative Genomics Viewer (IGV)-compatible .seg format and contains the sample name, chromosome, start and end position of the probe locus, probe region length, and probes tumor/normal log2 intensity ratio. The same file also contains B-nAllele frequency data ranging from zero to one for the tumor and matched normal sample.
2. ”SegData.seg” file containing copy number segment calls in IGV.seg format and output from GATK4.0 best practices pipeline.
3. “Variants.bed” file containing genomic positions and annotations for somatic and germline sequence-level variants as well as their tumor VAF, gene expression tpm values or similar for genes of interest in six-column bed format (https://genome.ucsc.edu/FAQ/FAQformat.html#format1)
4. “Fusions.bedpe” file containing fusion gene coordinates and annotations in bedpe format (https://bedtools.readthedocs.io/en/latest/content/general-usage.html#bedpe-format)

GISTIC bed files were generated by TCGA, Firehose Legacy pipeline^11^ and downloaded using the TCGAbiolinks package (https://rdrr.io/github/BioinformaticsFMRP/TCGAbiolinks/man/getGistic.html).

### Copy number probe and segment rescaling

Rescaling of the copy number probe and segment tracks is based on the normal copy number *n* = *ploidy* * (1 - *purity*), the tumor copy number *t* = *c* * *purity* and the corresponding log_2_ ratio *lrat* = log_2_((*t* + *n*) / *ploidy*), where *purity* is the purity estimate and *ploidy* is the ploidy estimate.

From this, the real tumor copy number *cnreal* can be obtained as

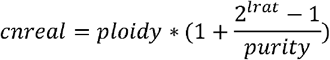

### BAF rescaling

The B-allele frequency (BAF) track shows the proportion of each alternative allele at SNP loci along the genome. We apply BAF correction according to the CNVkit ^12^ (https://cnvkit.readthedocs.io/en/stable/index.html).

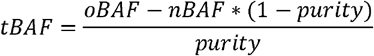

Where *tBAF* is the original tumor BAF, which is calculated based on the observed BAF *oBAF*, the normal BAF *nBAF* and the tumor purity *purity*.

### Statistical Analyses

We reported the time length for clinical case review with a rounded time to the nearest minute and summarized results using the median and interquartile range (IQR). We performed a normality test for two groups using Shapiro-Wilk normality test and used the Wilcoxon rank sum test to assess the time difference between our standard laboratory SOP and CNAEL for non-normally distributed data.

Statistical significance was defined as a two-sided *P*-value < 0.05, unless otherwise noted. All statistical analyses used R software V.4.2.1.

## Results

### Description of the software and web application

We developed and implemented CNAEL using Javascript and PHP with a MySQL backend database and deployed on an AWS elastic instance. The user can interact with the application through any web browser. A demonstration instance is available at https://CNAEL.sema4.com/demostration and example input data can be found at https://CNAEL.sema4.com/example. The peak RAM consumption from exome derived CNV data is approximately 32 GB and user interface renders in seconds. Input data are IGV-compatible seg files containing copy number segments and probe-level data (https://software.broadinstitute.org/software/igv/SEG). The software assumes the copy number call to be a tumor/normal log2 ratio, which can be generated from the GATK4.0 best practices somatic CNV detection pipeline (https://github.com/gatk-workflows/gatk4-somatic-cnvs), or an analogous software pipeline. Optionally, the user may upload somatic and germline variant calls in vcf format, gene expression values in tags per million (TPM) format, and fusion gene calls from tools such as STAR fusion. Source code along with example data is provided on (https://github.com/Sema4-Research/CNEAL). **Figure 1** shows an overview of the CNAEL user interface.

**Figure 1.**
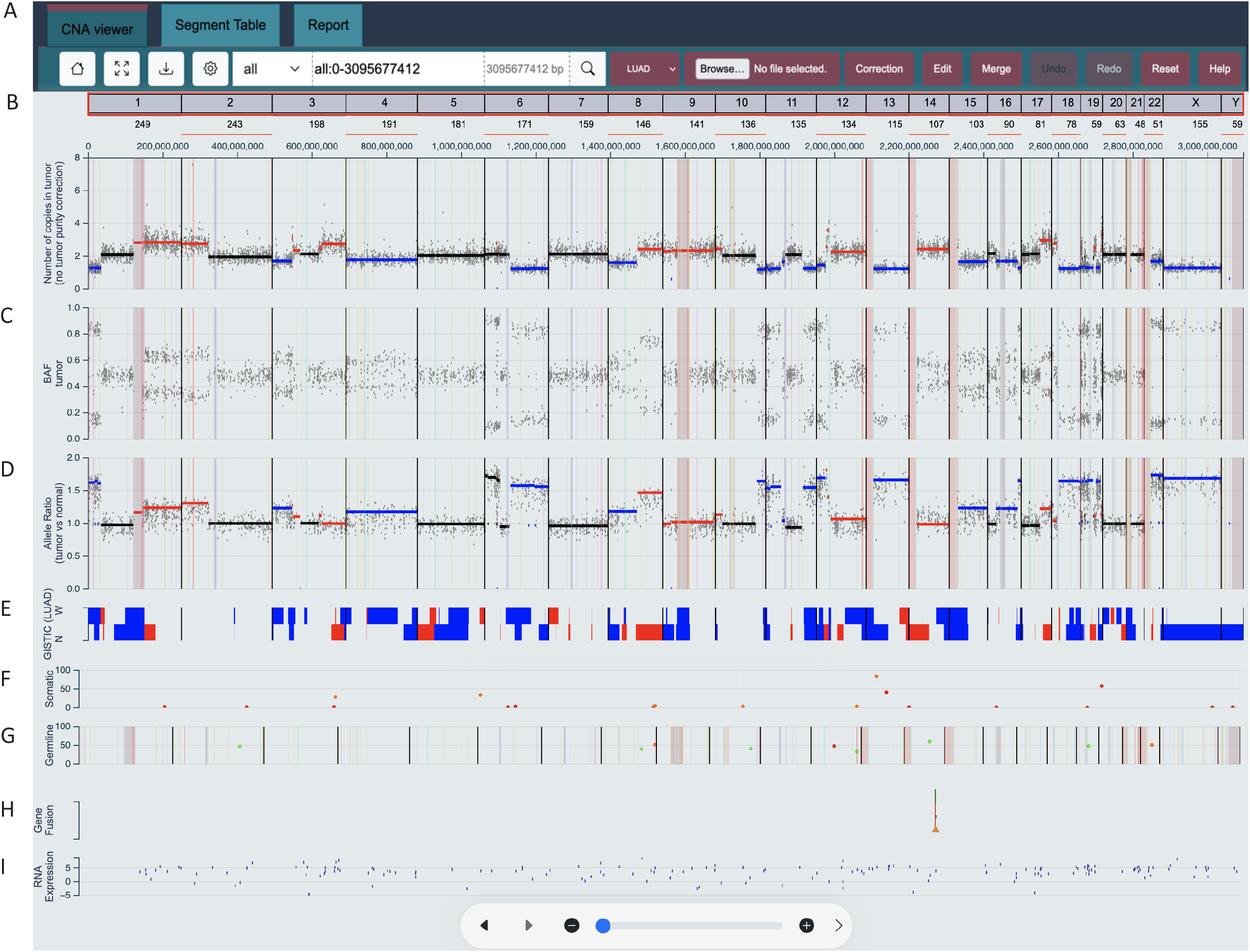
Overview of the CNA Explorer and Analyzer (CNAEL) graphical user interface (GUI). **A)** Navigation bar and other controls **B)** Copy number plot shows predicted copy number of each locus assayed: copy number segments are superimposed as horizontal bars with losses in blue, gains and amplifications in red and neutral in black. These plots may be rescaled based on tumor purity, ploidy and modal chromosome number by using the “correction” button, above the plot **C)** B-Allele Frequency (BAF) plot **D)** Allelic ratio plot. Gained/lost/amplified regions are colored corresponding to the copy number plot in (b) **E)** GISTIC plot shows wide (W) and narrow (N) segments predicted to be significantly enriched in a specific tumor type. Tumor type and corresponding GISTIC plots may be changed by clicking the OncoTree code box **F)** Somatic mutations, in this case called by Mutect2, with their genomic position on the x-axis and variant allele percentage on the y-axis **G)** Germline mutations in cancer-relevant genes. Note that the germline track also shows sequence assembly gaps and repeats loci as vertical red and grey bars – this feature is user-configurable and is switched off for the somatic track in this example. **H)** Predicted gene fusions from RNA-Seq. **I)** RNA-Seq expression of cancer relevant genes shown as log2(tags per million). Note that other facets of the GUI including interactive segment tables and genomic segment summarization are presented in **Supplementary Figure 2** and **Supplementary Figure 3**.

### SOP to operationalize ACMG/CGC guidelines

Technical review and reporting of exome-derived tumor copy number data are major bottlenecks for our clinical lab with analysis of complex cases typically taking hours to review ^13-15^. To replace the industry standard but time-consuming segment-by-segment analysis, we devised a five-step process for efficient review of genome-wide CNA profiles. The process consists of 1) understanding the purity, ploidy, present copy levels, and rescaling the data if appropriate; 2) identifying clinically significant focal abnormalities; 3) identifying clinically significant broad (chromosome arm and whole chromosome level) copy number abnormalities; 4) summarizing complex and chromothripsis regions; 5) auto-summarization of the whole copy number profile for optional reporting.

#### Step 1: Understand the purity, ploidy and copy levels

Approximately one third of tumors from pan-cancer studies undergo polyploidization, such as whole genome doubling ^16,17^. Polyploid states such as tetraploidy may be a prognostic indicator in certain tumor types ^16^ and correct assessment of ploidy is an important component of copy number analysis. However, many copy number segmentation algorithms do not consider tumor ploidy or purity when generating segment copy number ratios, whereas bioinformatic approaches such as PureCN and ASCAT generate multiple, and sometimes, equally likely purity/ploidy solutions ^18,19^. Furthermore, the modal (or ‘normal’) chromosome number is sometimes mis-identified by the algorithm, and these misconfigurations can lead to incorrect clinical interpretation. For example, 1p/19q co-deleted oligodendroglioma near tetraploid may be misinterpreted as having gains of every genomic region *except* for 1p and 19q, if the baseline copy number of two is selected by the segmentation algorithm (**Figure 2a**). Such deficiencies in bioinformatic segmentation necessitate either laborious manual correction of segments to reflect their true state or rerunning of the bioinformatics pipeline with user supplied parameters, which is operationally complex. We estimate that 10-30% of tumors from our clinical service are subject to these issues and require manual intervention. An example of a tumor with an incorrect copy number baseline and its subsequent correction is shown in **Figure 2b-c**.

**Figure 2.**
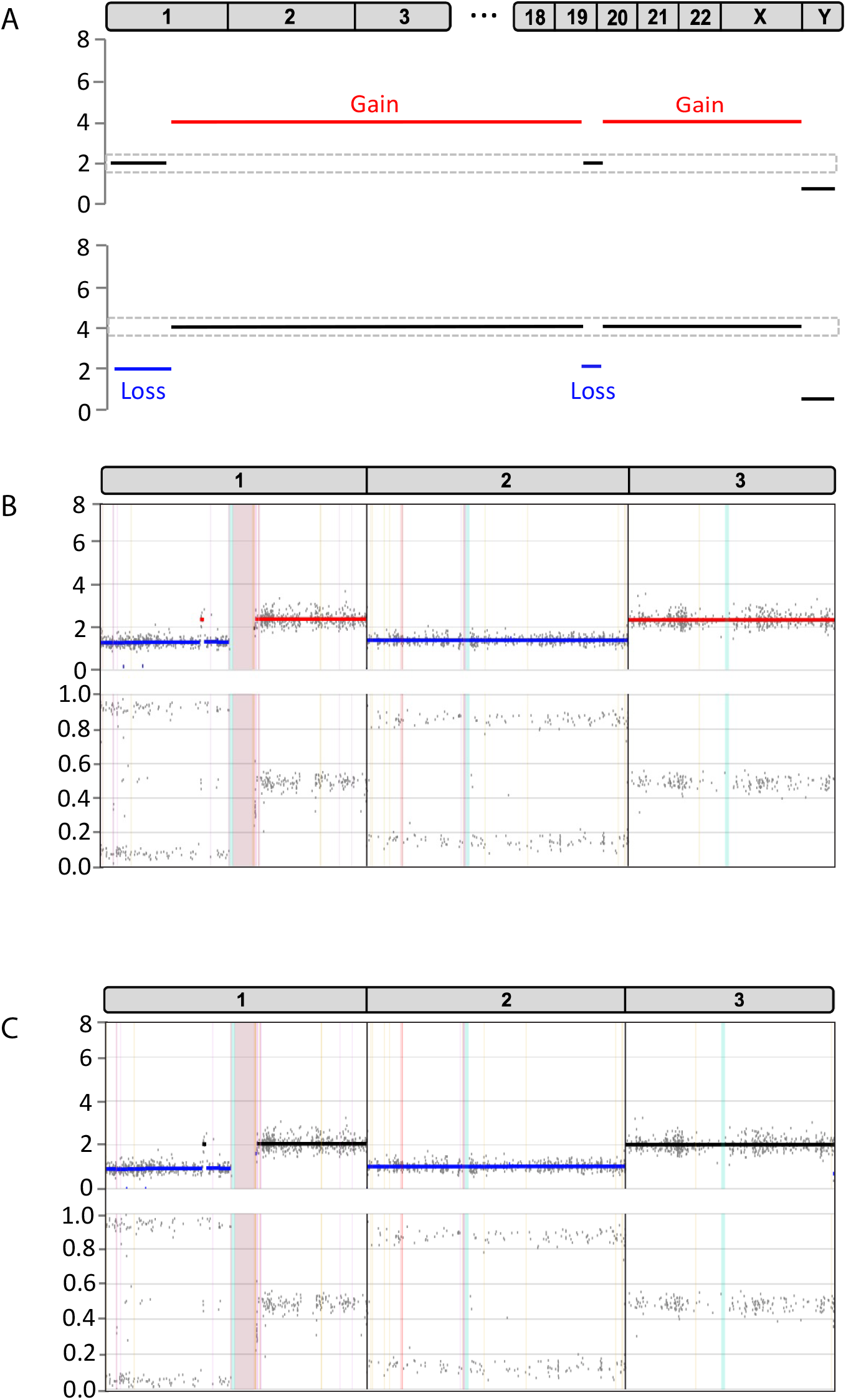
Effects of incorrect copy number baseline on interpretation. **A)** Upper: Schematic representation of a 1p/19q co-deletion in a near tetraploid tumor genome. Grey boxes on the x-axis represent chromosomes 1-3, 18-22, X, and Y with intervening chromosomes not shown for simplicity. Segment copy numbers are on the y-axis. Segments are called as gained (red), and neutral (black) when a baseline copy number of two (grey hashed box) is applied to the case. Lower: When the modal chromosome number of four is used as a baseline, losses of 1p and 19q (blue) are now evident, leading to the correct interpretation of the case: 1p/19q co-deletion. **B)** An example from our clinical lab shows an incorrect copy number baseline has been selected by GATK4.0 resulting in gain calls for 1q and the whole of chromosome 3. These require an analyst to review and correct to ‘false positive’ calls or else an erroneous copy number report may be issued. **C)** Rescaling of the data using CNAEL shifts the erroneously called segments down to a neutral state: the correct clinical interpretation for this case.

CNAEL deploys a novel rescaling algorithm that allows users to convert a segment’s tumor/normal log2 ratio, as is output by most CNA segmentation algorithms, to a real segment copy number while accounting for user-supplied tumor purity, ploidy, and modal chromosome number (**Materials and Methods**). Purity and ploidy estimates can come from external data such as anatomic pathology reports, FISH, algorithms such as PureCN, or be derived from the data itself by interpreting copy levels present in the context of and allelic imbalance /BAFs present in the sample. **Supplementary Figure 1** shows further examples of our on-the-fly copy number correction of polyploid and low tumor purity samples.

#### Step 2: Identify clinically significant focal abnormalities

Once the copy number baseline has been established, the analyst can identify focal amplifications and deletions containing oncogenes or tumor suppressor genes, respectively. ‘Focal’ lacks a strict definition, while in our SOP, we considered segments of less than 5 Mb to be potentially focal. Similarly, according to official guidelines, ‘amplification’ is often defined in a disease-specific manner. For simplicity, we defined it here as >= 3X the modal chromosome number. Primarily, focal abnormalities can be reviewed in a disease-agnostic manner. For example, a focal amplification of *ERRB2* accompanied by RNA overexpression would likely be deemed clinically significant and potentially targetable, regardless of tumor type (although of specific significance in breast cancer).

CNAEL facilitates rapid identification of gain, amplification, and deletion target genes in four ways: a) a user-configurable threshold for a segment to be called “amplified” as opposed to “gained” for example, six or more copies in a diploid genome versus twelve or more copies in a tetraploid one to be called an amplification; b) a filterable and sortable tabular view of copy number segments that includes gene content, oncogene/tumor suppressor gene status and knowledgebase hits; c) an interactive multi-variant visualization that highlights sequence-level somatic or germline mutations facilitating identification of oncogenic gain of function mutations or tumor suppressors with two hits (**Figure 3a-g**); d) If RNA-Seq counts data is supplied, outlier expression of cancer-relevant genes will be displayed to help identify gain or amplification target genes within regions of complex amplification (**Figure 3f**). Once significant segments have been identified, they can be classified as clinically significant (Tier 1 or 2, according to guidelines) and reported.

**Figure 3.**
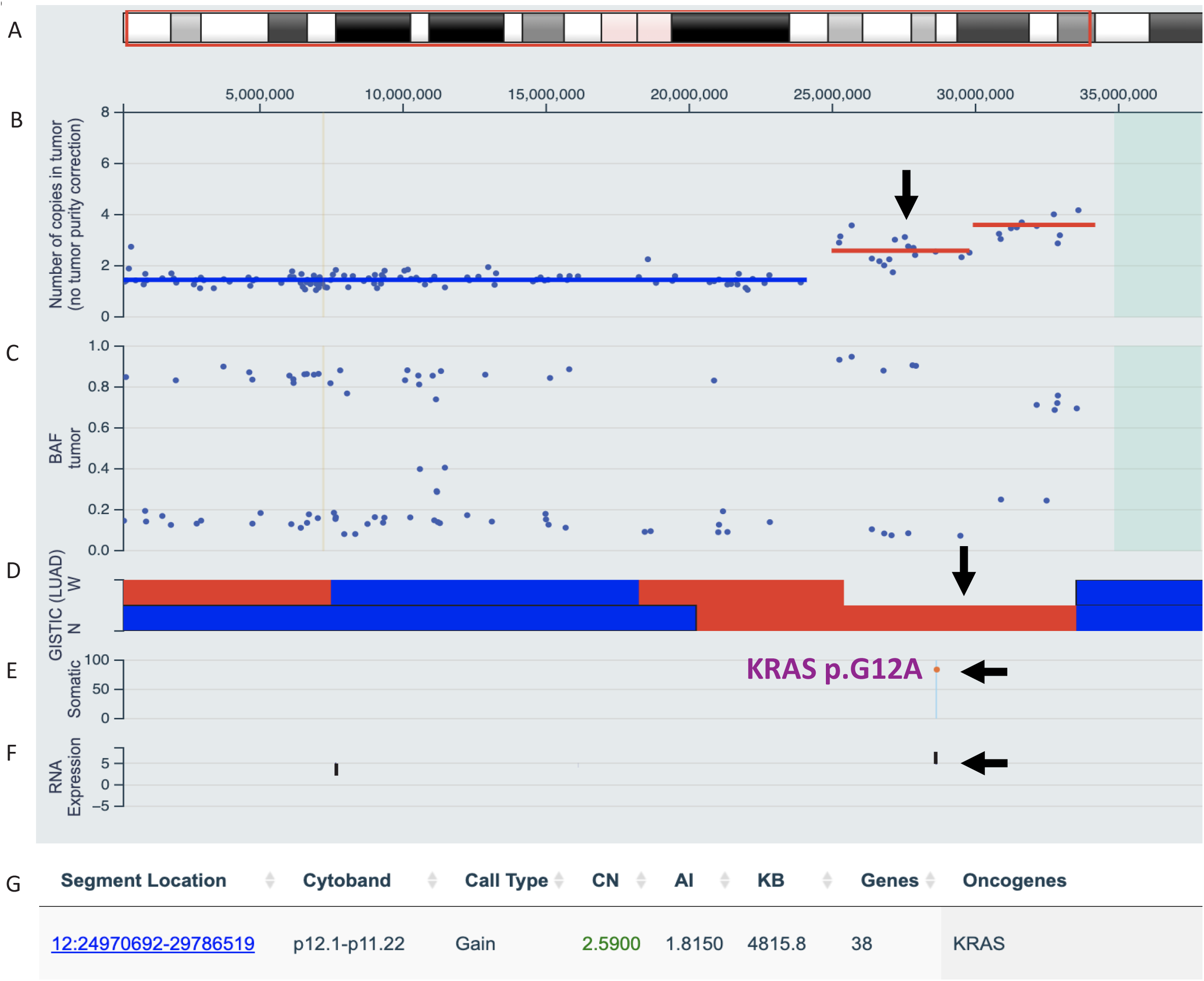
Identifying clinically significant focal CNAs; Chromosome 12p of a lung adenocarcinoma (LUAD). **A)** Chromosome ideogram showing cytobands. The region depicted is chr12:1-38,000,000[hg19]. **B)** Called copy number segments (blue for loss and red for gain) and exon probe intensities (scatter plot) shows three called segments on 12p. **C)** B-Allele frequency supports a copy number imbalance along the whole of 12p. **D)** LUAD GISTIC tracks show that losses (blue) and gains (red) of 12p are found recurrently in LUAD. Regions from the GISTIC “Narrow peak” configuration are labeled as “N” and “wide” as “W”. As the loss of distal 12p is recurrent, it should be assessed further for specific clinical significance. Proximal 12q shows a narrow GISTIC region that overlaps with the two gained segments in this case (black arrow). **E)** A somatic mutation of *KRAS* (p.G12A) with a variant allele frequency of 0.84 overlaps the first gained segment suggesting that the mutant allele is in a gained state. **F)** RNA expression of *KRAS* is elevated in this case with a TPM of 84 suggesting that the gain may be driving high expression. **G)** CNAEL tabular view highlights *KRAS* as the likely driver of this copy number gain (CN = Copy number, AI = Allelic imbalance, KB = kilo bases). In conclusion, distal 12p loss is a recurrent abnormality found in this tumor type but with no additional clinical significance (tier 3) and can be reported in the copy number appendix, chr12:24970692-29786519 is a clinically significant (tier 2) one copy as it bears a *KRAS* hotspot mutation (black arrow, panel A). Inhibitors downstream of *KRAS* may be a therapeutic option.

#### Step 3: Identify clinically significant broad copy number abnormalities including co-occurring abnormalities

Broad copy number abnormalities are the most challenging to interpret because their clinical significance is often disease specific. For example, deletion of chromosome 16q is recurrent in breast cancer, which may have sufficient evidence to report it as a tier 2 abnormality ^20^. In other tumor types such as ovarian cancer, assuming no double hits are present, there is less evidence of significance, making this abnormality a tier 3 event and is less likely to be reported ^21^. To aid analysts in identifying recurrent broad copy number abnormalities that may be of clinical significance, CNAEL provides a tumor type-specific Genomic Identification of Significant Targets in Cancer (GISTIC) track that is derived from public data ^11,22^. The user may then identify CNA regions with significant overlap to an OncoTree-specific GISTIC plot (**Supplementary Figure 4**). Those regions with overlap are likely enriched above the level of statistical noise for that tumor type and become candidates for further investigation for therapeutic or prognostic association and subsequent reporting as tier 1 or 2 abnormalities. Currently, segment overlap between the GISTIC regions and present tumor segments is assessed visually, but there could be an opportunity for automation in the future.

**Figure 4.**
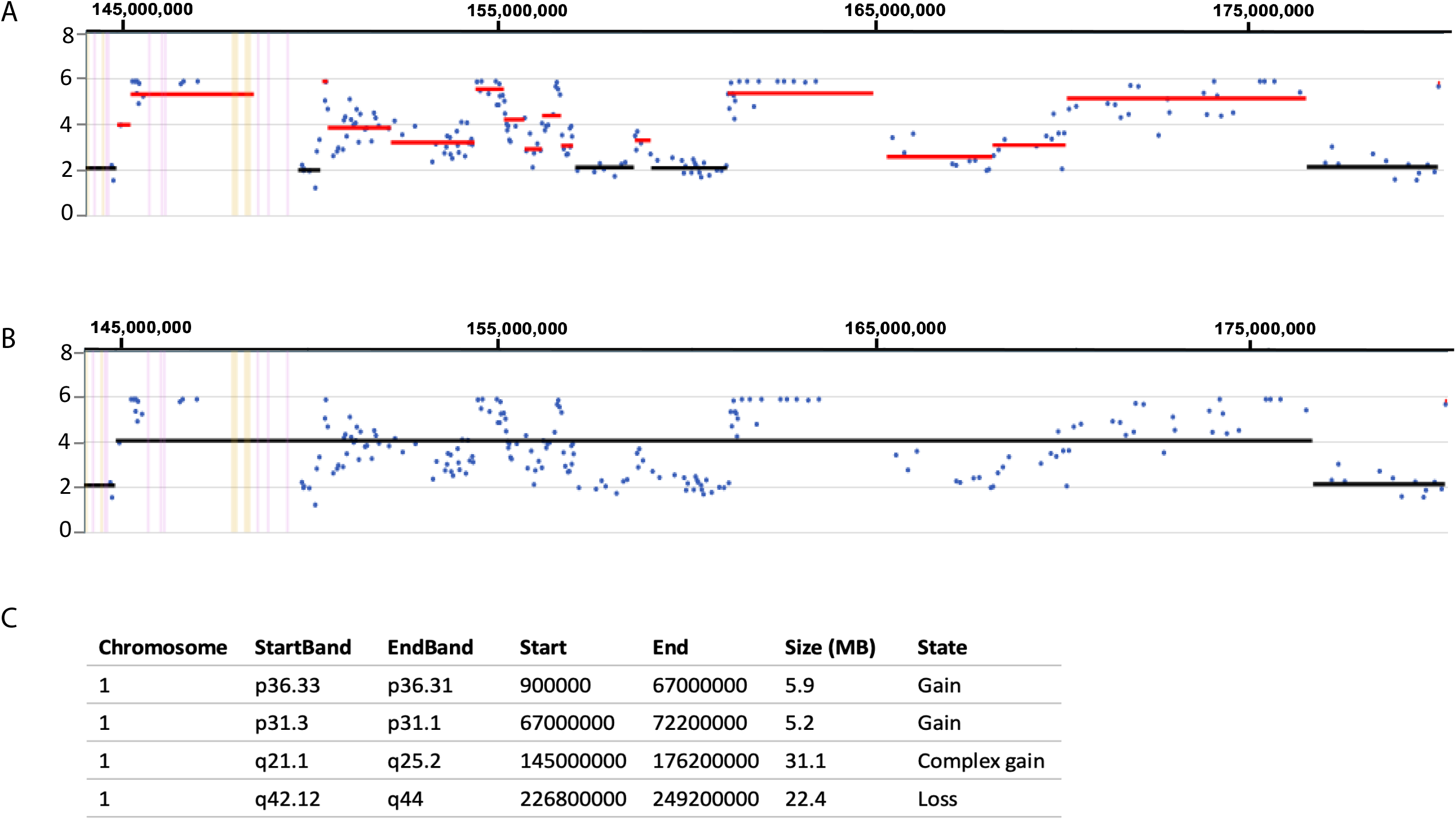
Collapsing a complex region to a single call. **A)** Copy number probes and segments of an approximately 30 Mb region of chromosome 1q in a prostate cancer case. This tumor has a complex series of gains totaling fifteen distinct copy number segments. A clear genetic driver of this complex rearrangement could not be identified. **B)** Collapsing this region to a single “complex gain” encompassing approximately 145-176 Mb. **C)** Automated summary table shows the complex gain among other called segments on chromosome 1 at 5 Mb resolution and above.

#### Step 4: Summarize complex and chromothripsis regions

Tumor genomes can contain regions of complex copy number where numerous losses, gains or amplifications are found on a single chromosome or chromosome arm, but a clear driver gene cannot be identified. Further, chromosome shattering and reassembly due to chromothripsis can produce hundreds of copy number segments in certain cases ^23^. Reporting such regions is a quandary as they are clearly the result of an oncogenic process but if reported, the many segments of varying copy number make interpretation challenging for the clinician. CNAEL operationalizes guideline recommendations to summarize these complex and chromothripsis regions into a single call by allowing the user to collapse such regions into a single ‘complex’ or chromothripsis’ call to simplify reporting (**Figure 4a-b**). The net result is a succinct report that notes a complex region resulting from an oncogenic process. At present, the user must manually identify the segments to be collapsed into a single call, but automated detection of candidate complex regions may be possible through machine learning in the future.

#### Step 5: Summarize the remaining segments

A description of the abnormal tumor copy number profile is often desirable even if no segments are found to be of tier 1 or 2 clinical significance. Such descriptions are useful in ascertaining if tumor tissue was present in the provided specimen, for comparison with prior or subsequent samples from the same individual or to facilitate literature searches. Not reporting these tier 3 abnormalities may create the erroneous impression of a ‘normal’ copy number profile. In review of our previous SOP, we found that analysts were spending an excessive amount of time manually correcting segment start and end positions and merging bioinformatically over-segmented regions to generate a ‘copy number appendix’ even though these segments have no known clinical actionability. CNAEL takes an automated approach to profile summarization by merging segments > 5 Mb into contiguous loss/gain/amplification calls relative to the modal chromosome number without the analyst having to perform any hands-on operations. CNAEL does have an ability to manually create, edit, and merge segments, but our recommendation is to use this feature only for tier 1 and 2 variants because their precise description for reporting is vital, whereas a lower-resolution summarized description of the genome is acceptable for the copy number appendix (**Figure 4c**).

### Hands on analysis time

We selected twenty moderate to high-complexity cases from our clinical service representing 17 distinct tumor types. These samples had undergone tumor/normal exome sequencing and copy number segments were called with a GATK4.0 best practices pipeline. The total number of called copy number segments ranged from 27 to 261 with a median of 88 and copy number profiles contained a variety of focal, broad, and tier 1-3 abnormalities. Two separate sets of analysts reviewed cases prior to lab director approval using i) a segment-by-segment assessment of gene content and manual correction of segment copy number calls ii) using CNAEL and the associated SOP. Both sets of analysts blindly reviewed the cases. We performed normality test for both review time of CNAEL (p= 0.2709) and standard SOP (p = 0.0067). In every case among all 20 cases, we identified a significantly faster analysis turnaround time using CNAEL with a median of 7 mins [IQR = 4, 10.25] compared with the standard laboratory SOP we followed with a median of 61 mins [IQR = 23.75, 176,25], with p=4.631e-05 by Wilcoxon rank sum test. This is a 9 fold decrease in review time and the review resulted in a clinically equivalent report being issued as assessed by an American College of Medical Genetics and Genomics-certified laboratory director (**Table 1**).

**Table 1.** Clinical case review time length (minutes) by our standard SOP (Segment by segment) and CNAEL. The table lists case 1-case 20 for 17 distinct cancers (oncotree codes), a total number of called segments per case (Number of Called Segments), review time by our standard SOP (Segment by segment), and CNAEL. Our standard SOP time is counted from curator starting the review of the CNA segment to submitting segments followed by our SOP. The review time period was automatically recorded by our computational curation system. CNAEL time is counted the total minutes spent on the same CNA segments curated for each case.

| Case ID | OncoTree | Number Called Segments | Segment by segment review time (minutes) | CNAEL time (minutes) |
| --- | --- | --- | --- | --- |
| Case_1 | LGFMS | 52 | 33 | 7 |
| Case_2 | USC | 150 | 381 | 9 |
| Case_3 | ULMS | 171 | 110 | 11 |
| Case_4 | PRAD | 132 | 219 | 15 |
| Case_5 | CHS | 65 | 36 | 7 |
| Case_6 | MACR | 49 | 34 | 4 |
| Case_7 | PRCC | 39 | 88 | 5 |
| Case_8 | NETNOS | 77 | 45 | 8 |
| Case_9 | PRAD | 167 | 186 | 15 |
| Case_10 | STAD | 36 | 1 | 1 |
| Case_11 | LUAD | 143 | 5 | 2 |
| Case_12 | TPLL | 105 | 260 | 17 |
| Case_13 | MACR | 43 | 9 | 10 |
| Case_14 | PANET | 133 | 250 | 14 |
| Case_15 | OOVC | 261 | 8 | 5 |
| Case_16 | PAST | 27 | 17 | 6 |
| Case_17 | LUAD | 162 | 173 | 10 |
| Case_18 | PCM | 46 | 121 | 3 |
| Case_19 | ISTAD | 98 | 26 | 3 |
| Case_20 | ARMS | 65 | 77 | 4 |

## Discussion

Tumor CNA profiles contain a wealth of information that can inform clinical actionability. However, review and reporting procedures remain complex despite recent progress made on standardization through the consensus guidelines ^9^. As a result, many popular tumor NGS tests underutilize CNA profiles and focus solely on focal amplification and deep (likely homozygous) deletion of well-established oncogenes and tumor suppressor genes ^11,24^. Clinical laboratories that comprehensively report CNAs frequently experience a bottleneck for such analysis, resulting in a slow case turnaround and an inefficient use of personnel^13-15^.

Our group took two approaches to address these issues: Firstly, we broke down the consensus guidelines for CNA reporting into five discrete steps that can be operationalized through an efficient analysis SOP. These steps are generic enough that any clinical laboratory wishing to perform genome-wide copy number analysis can use them. Secondly, we designed an interactive web application that alleviates common review bottlenecks. For example, by removing the need to rerun a bioinformatics pipeline with user-supplied parameters defining purity, ploidy, and modal chromosome number and integrating mutational and gene expression data when available. The SOP combined with the software application results in a significantly reduced analysis turnaround time for a small series of cases.

Similar open-source software solutions exist, such as ReconCNV ^25^ and ShinyCNV ^26^ as well as paid software including, Biodiscovery Nexus and Thermofisher Chromosome Analysis Suite. However, to our knowledge – none of these softwares is as fully featured as CNAEL and lacks the ability to dynamically re-scale data, summarize complex regions, perform a multi-platform analysis, or facilitate tight integrations with official CNA analysis guidelines.

In future versions of CNAEL, we wish to further optimize the analysis SOP by employing automated approaches to detect complex and chromothripsis regions and identify clinically significant co-occurring abnormalities, such as hyperdiploidy and 1p/19q co-deletion, in their specific tumor types. Better representation of the clonal or subclonal nature of the CNAs after adjusting for tumor ploidy and purity may also benefit clinical reporting. Lastly, genome wide estimates of chromosome instability and LOH might also be implemented as such abnormalities have been linked to prognosis and to response to PARP inhibitors in certain tumor types ^27,28^.

In conclusion, CNAEL and our analysis SOP reduces the complexity of tumor CNA analysis. These steps outlined in this study can be used to optimize clinical sequencing operations.

## Supporting information

SupFig 1

SupFig 2

SupFig 3

SupFig 4

## Data Availability

CNAEL is available at https://CNAEL.sema4.com ; Source code and data from the 20 cases analyzed as part of this study is available at https://github.com/Sema4-Research/CNEAL.

## Acknowledgements

We also thank Sema4 IT and health informatics staff Shay Hassidim, Indra Harahap, Caleb Chandrabose, and Michael Distefano for supporting our AWS computing and needs.

## Author Information

S.N., H.L. designed the study. E.C., H.L. designed the framework of the user interface. H.A.K, A.S and S.N. developed the optimized analysis SOP. E.C., J.W. wrote code and developed the web application. R.K. derived data rescaling equations. L.L., A.U. wrote bioinformatics pipelines upstream of the web application. A.S and S.N. performed clinical case review. J.W., S.N., and E.C. wrote the manuscript. All authors participated in the data interpretation, manuscript editing, and approval of the final submitted version.

## Ethics Declaration

Ethics approval was not required for our study as it uses de-identfied data under the HIPAA privacy rule.

## Legends

**Supplementary Figure 1. Additional Examples of data rescaling based on tumor purity and ploidy**. Upper plots show the original copy number segmentation output from GATK4, and lower plots show the corrected and rescaled plots **A)** Triploid sample, 90% purity. **B)** Triploid sample, 50% purity. **C)** Tetraploid sample, 100% purity.

**Supplementary Figure 2. Segment table component of the GUI**. The second facet of the CNAEL GUI is an interactive copy number segments table. CN = Copy number, AI = Allelic imbalance, KB = kilo bases, CAV = Gene matches to our internal knowledgebase. CN values will update automatically if the user rescales the data using the copy number correction function. Oncogene and tumor suppressor gene annotations are based on literature review by our knowledgebase team.

**Supplementary Figure 3. Reporting component of the GUI**. The third facet of the CNAEL GUI is a reporting page. Users may write a case summary in the free form text box. Segments designated as “Clinically Significant” will appear in the middle section and will appear at the top of the report. The copy number appendix is built automatically by the CNAEL software to save the user time in compiling a genome-wide list of copy number segments for inclusion in the copy number report as part of the “Copy Number Appendix.” The underlying code will automatically merge segments of the same designation (loss/gain/amplification) and report any region of > 5 Mb.

**Supplementary Figure 4. GISTIC Overlaps of chromosome 5 in an AML case**. Chromosome 5 of an AML case shows a one-copy interstitial deletion of chromosome 5q (chr5:99921635-147593838[hg19]; upper plot (blue segment) is accompanied by a split in the B-Allele frequency. Both narrow and broad GISTIC tracks (lower blue ribbon plots) overlap the deleted region, which alerts the user of a potential clinical significance.

