## Supplementary figures and images for "CNA Explorer and anaLyzer (CNAEL): an interactive web application and standard operating procedure enabling efficient clinical review and reporting of complex NGS-derived tumor copy number profiles"

### SupFig 1

#### Supplemental Figure 1

## 1A

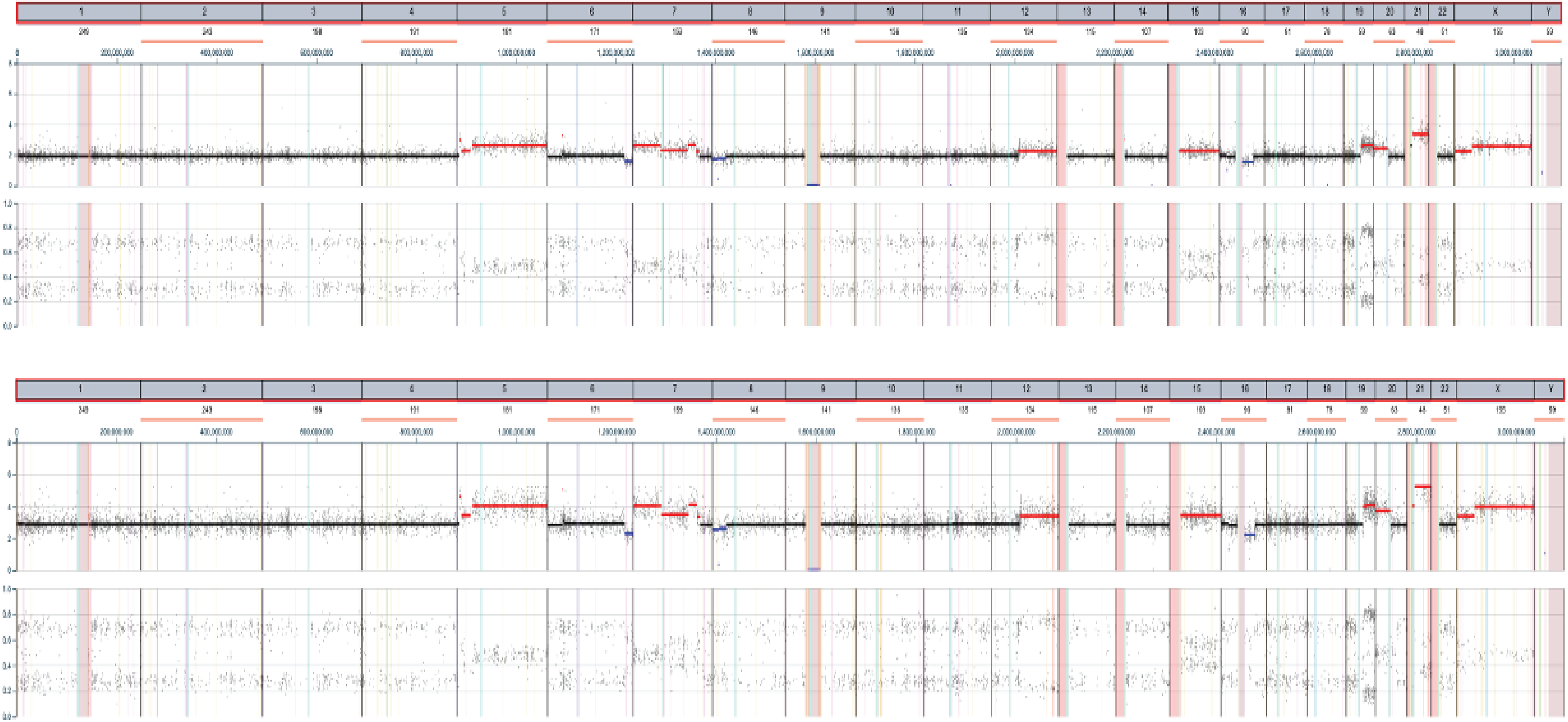

**1B**

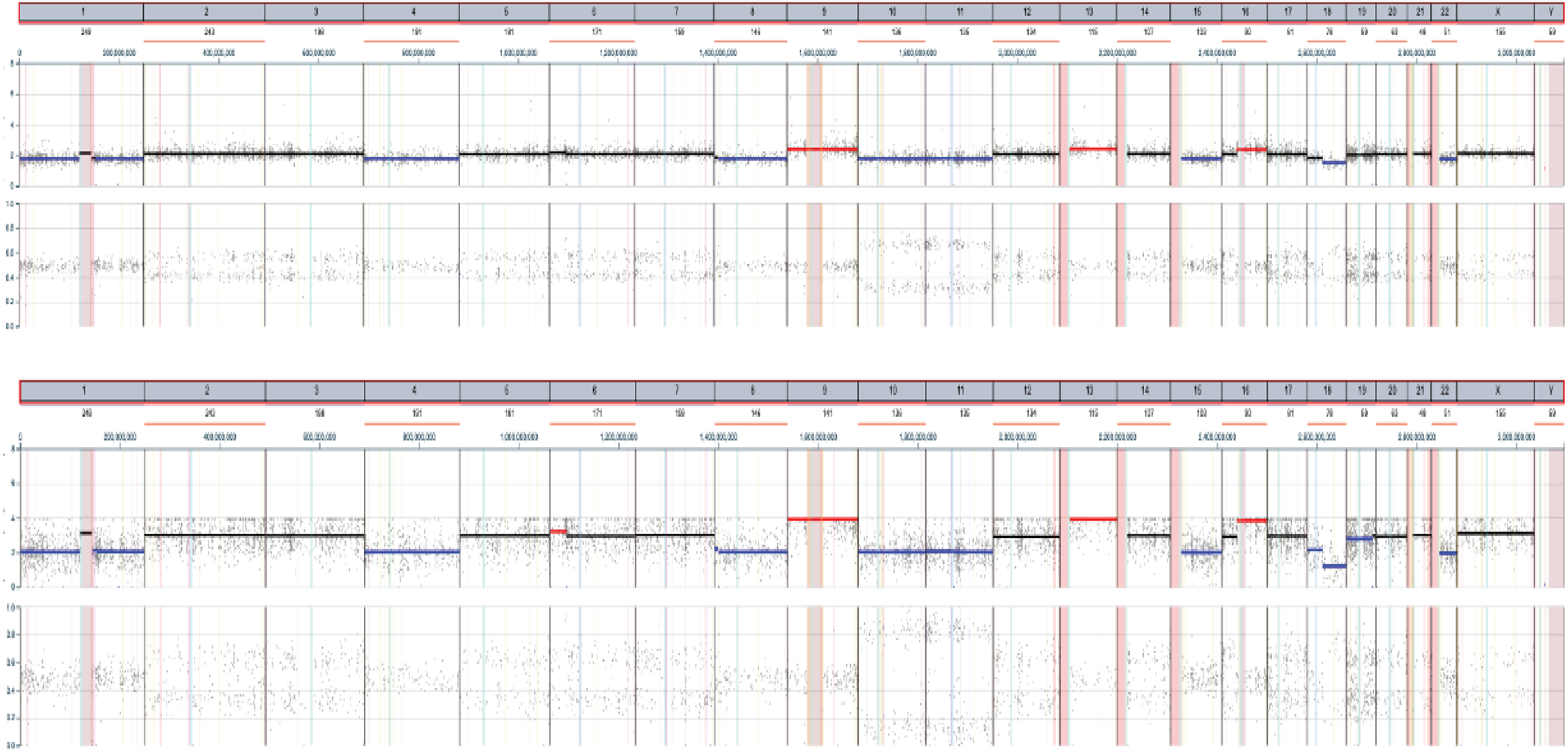

**1C**

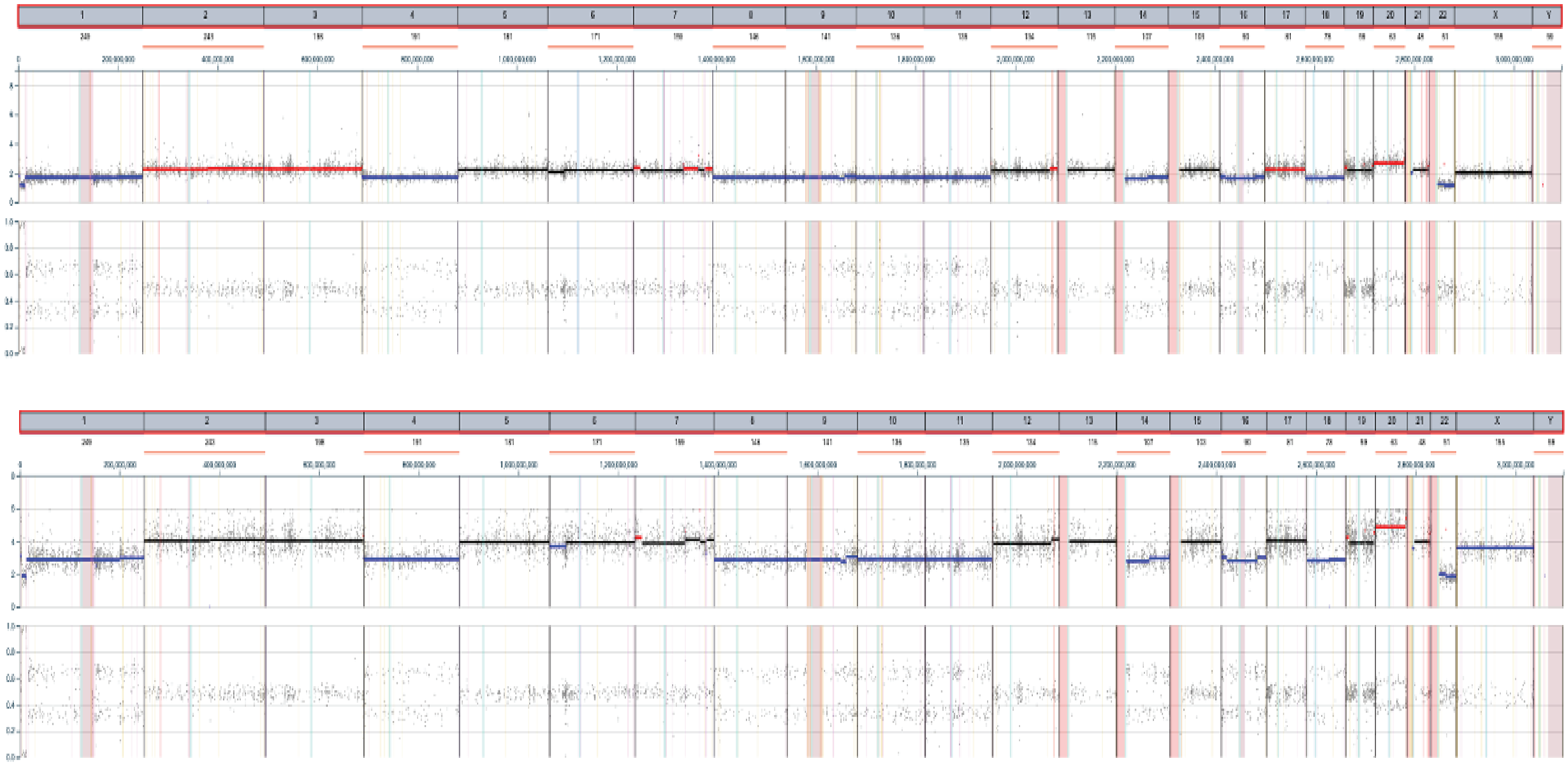

### SupFig 4

##### Supplemental Figure 4

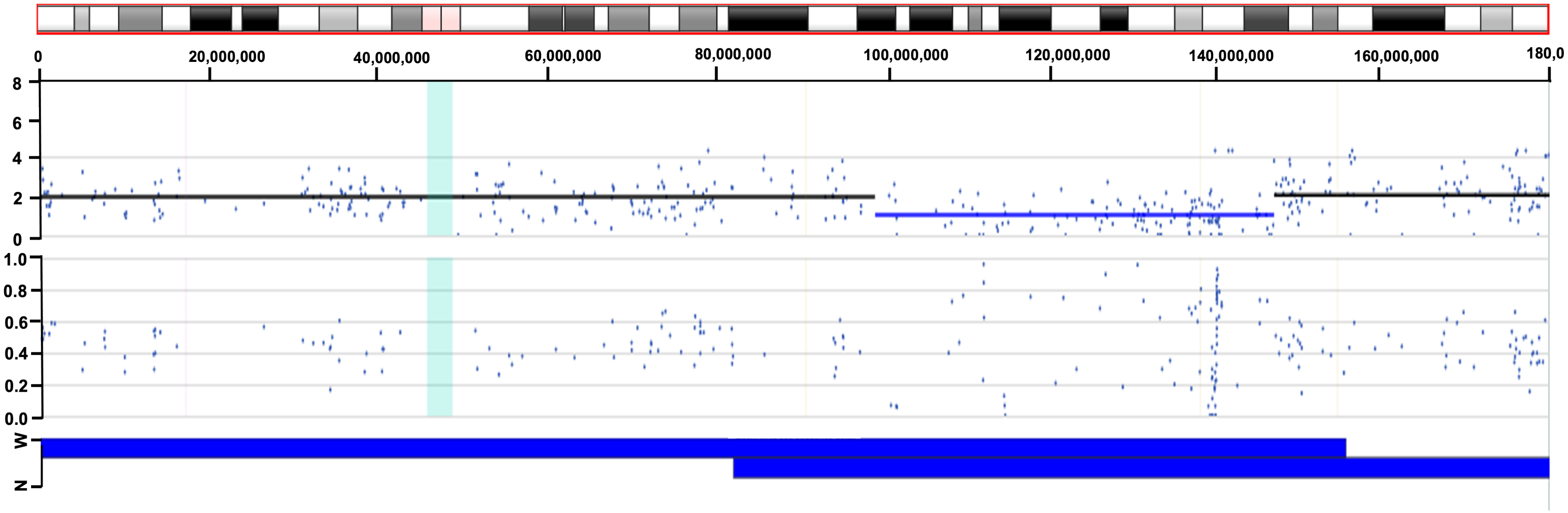
