## Supplementary material for "CNA Explorer and anaLyzer (CNAEL): an interactive web application and standard operating procedure enabling efficient clinical review and reporting of complex NGS-derived tumor copy number profiles": SupFig 2

### Supplemental Figure 2

Show 25 entries ☑ Appendix only ☑ CS ☑ VUS ☑ Benign ☑ False positive Search:

| Segment Location | Cytoband | Call Type | CN | AI | KB | Genes | Oncogenes | Tumor Suppressor Gene | CAV | Notes | Clin Sig |
|---|---|---|---|---|---|---|---|---|---|---|---|
| 10:111624692-135439267 | q25.1-q26.3 | Loss | 1.2000 | 1.6434 | 23814.6 | 205 | ADRB1 BUB3 FGFR2 GRK5 MKI67 SHOC2 TCF7L2 | BUB3 GRK5 MGMT SMC3 | | | Appendix only |
| 10:18439562-108924476 | p12.33-q25.1 | normal | 2.0400 | 0.9939 | 90484.9 | 750 | A1CF ABCC2 ACTA2 BMI1 BMPR1A BTRC CDK1 CREM CYP17A1 ERCC6 FGF8 GOT1 HELLS JMJD1C LDB1 MAP3K8 MAPK8 MLLT10 NFKB2 NRG3 NT5C2 PAX2 PDCD11 RASGEF1A RET SIRT1 TET1 TLX1 TNKS2 UBE2D1 WNT8B ZEB1 | ABI1 ALOX5 ARID5B BLNK BMPR1A BTRC CACNB2 CCDC6 CPEB3 CREM CTNNA3 CYP2C8 EGR2 FAS IFIT1 IFIT2 IFIT3 KAT6B KLLN NCOA4 NFKB2 NKX2-3 PRF1 PTEN SIRT1 SUFU TET1 | | | Appendix only |
| 10:92747-18429947 | p15.3-p12.33 | Gain | 2.4400 | 1.1340 | 18337.2 | 153 | GATA3 GTPBP4 IL2RA SUV39H2 | CACNB2 GATA3 IL15RA KLF6 LARP4B ZMYND11 | | | Appendix only |
| 11:12183453-12229936 | p15.3-p15.3 | Loss | 1.1400 | 0.9518 | 46.5 | 2 | | | | | Appendix only |
| 11:12230794-40138092 | p15.3-p12 | Loss | 1.2500 | 1.5587 | 27907.3 | 180 | BDNF EHF ELF5 LGR4 LMO2 MYOD1 PAX6 PIK3C2A PRMT3 TEAD1 TRAF6 WT1 | CD44 CYP2R1 ELF5 FANCF WT1 | | | Appendix only |
| 11:192850-197487 | p15.5-p15.5 | Loss | 1.4800 | 0.9789 | 4.6 | 3 | | | | | Appendix only |
| 11:197488-5863377 | p15.5-p15.4 | Loss | 1.3000 | 1.5394 | 5665.9 | 199 | ASCL2 CTSD H19 HRAS IGF2 NUP98 RRM1 | CARS1 CDKN1C H19 KCNQ1 MUC6 RRM1 | | | Appendix only |
