## Supplementary material for "CNA Explorer and anaLyzer (CNAEL): an interactive web application and standard operating procedure enabling efficient clinical review and reporting of complex NGS-derived tumor copy number profiles": SupFig 3

### Supplemental Figure 3

Save Export report

#### Summary

#### Clinical Significant segments

| Segment Location | Cytoband | Call Type | CN | AI | KB | Notes | Clin Sign |
|---|---|---|---|---|---|---|---|

#### Appendix

| Chromosome | StartBand | EndBand | Start | End | Size (MB) | State |
|---|---|---|---|---|---|---|
| 1 | p36.33 | p35.1 | 900000 | 33200000 | 32.3 | Loss |
| 1 | p11.2 | q44 | 121100000 | 249200000 | 128.1 | Gain |
| 2 | p25.3 | p13.2 | 0 | 71900000 | 71.9 | Gain |
| 3 | p26.3 | p21.1 | 400000 | 53400000 | 53.0 | Loss |
| 3 | p21.1 | p12.3 | 53500000 | 74500000 | 21.0 | Gain |
| 3 | q21.2 | q29 | 125600000 | 197800000 | 72.1 | Gain |
| 4 | p16.3 | q35.2 | 100000 | 190900000 | 190.8 | Loss |
| 6 | q12 | q27 | 69300000 | 170900000 | 101.5 | Loss |
| 8 | p23.3 | q21.11 | 200000 | 77900000 | 77.7 | Loss |
| 8 | q21.12 | q24.3 | 79500000 | 146300000 | 66.8 | Gain |
